# SARS-CoV-2 seroprevalence survey among 18,000 healthcare and administrative personnel at hospitals, pre-hospital services, and specialist practitioners in the Central Denmark Region

**DOI:** 10.1101/2020.08.10.20171850

**Authors:** Sanne Jespersen, Susan Mikkelsen, Thomas Greve, Kathrine Agergård Kaspersen, Martin Tolstrup, Jens Kjærgaard Boldsen, Jacob Dvinge Redder, Kent Nielsen, Anders Mønsted Abildgaard, Henrik Albert Kolstad, Lars Østergaard, Marianne Kragh Thomsen, Holger Jon Møller, Christian Erikstrup

## Abstract

**Objectives:** The objective of this study was to perform a large seroprevalence survey on severe acute respiratory syndrome coronavirus 2 (SARS-CoV-2) among Danish healthcare workers to identify high risk groups.

**Design:** Cross-sectional survey.

**Setting:** All healthcare workers and administrative personnel at the seven hospitals, pre-hospital services and specialist practitioner clinics in the Central Denmark Region were invited by e-mail to be tested for antibodies against SARS-CoV-2 by a commercial SARS-CoV-2 total antibody enzyme-linked immunosorbent assay (ELISA, Wantai Biological Pharmacy Enterprise Co., Ltd., Beijing, China).

**Participants:** A total of 25,950 participants were invited. Of these, 17,987 (69%) showed up for blood sampling, and 17,971 had samples available for SARS-CoV-2 antibody testing.

**Main outcome measures:** 1) Prevalence of SARS-CoV-2 antibodies; 2) Risk factors for seropositivity; 3) Association of SARS-CoV-2 RNA and antibodies.

**Results:** After adjustment for assay sensitivity and specificity, the overall seroprevalence was 3.4% (CI: 2.5%-3.8%). The seroprevalence was higher in the western part of the region than in the eastern part (11.9% vs 1.2%, difference: 10.7 percentage points, CI: 9.5-12.2). In the high prevalence area, the emergency departments had the highest seroprevalence (29.7%) while departments without patients or with limited patient contact had the lowest seroprevalence (2.2%). Multivariable logistic regression analysis with age, sex, and profession as the predictors showed that nursing staff, medical doctors, and biomedical laboratory scientists had a higher risk than medical secretaries, who served as reference (OR = 7.3, CI: 3.5–14.9; OR = 4., CI: 1.8–8.9; and OR = 5.0, CI: 2.1–11.6, respectively).

Among the total 668 seropositive participants, 433 (64.8%) had previously been tested for SARS-CoV-2 RNA, and 50.0% had a positive RT-PCR result. A total of 98% of individuals who had a previous positive viral RNA test were also found to be seropositive.

**Conclusions:** We found large differences in the prevalence of SARS-CoV-2 antibodies in staff working in the healthcare sector within a small geographical area of Denmark and signs of in-hospital transmission. Half of all seropositive staff had been tested positive by PCR prior to this survey. This study raises awareness of precautions which should be taken to avoid in-hospital transmission. Additionally, regular testing of healthcare workers for SARS-CoV-2 should be considered to identify areas with increased transmission.

**Trial registration:** The study is approved by the Danish Data Protection Agency (1-16-02-207-20).

## Introduction

During the year 2020, a pandemic caused by severe acute respiratory syndrome coronavirus 2 (SARS-CoV-2) has affected most countries in the world. However, the pandemic has not affected all countries or areas evenly. Thus, even in relatively small areas large differences in incidence rates have been observed (1). To mitigate the effects of the pandemic, health authorities have introduced interventions, e.g. the closing of schools, public institutions, prohibition of group gatherings, and even curfew. Healthcare workers may be at increased risk of infection (1, 2), but differences in seroprevalence according to professional status and use of personal protective equipment are present (1, 3, 4). Prevention of infection in healthcare workers is important not only to reduce morbidity and mortality in this population, but also to avoid secondary transmission and maintain the capacity of the healthcare system.

The objective of this study was to perform a seroprevalence survey among all healthcare and administrative personnel at hospitals, pre-hospital services, and specialist practitioners in the Central Denmark Region in order to identify high risk groups employed in the healthcare system, to find hotspots in the region, and to clarify whether the precautions for the healthcare professionals are sufficient. The survey was requested by the Danish Health Authority and the Danish Administrative Regions as a quality assurance project. Additionally, serological test results were compared with available results from previous SARS-CoV-2 RNA tests.

This study population is, to our knowledge, one of the largest in the world to date, demonstrating SARS-CoV-2 antibody screening among healthcare and administrative personnel. Indeed, the study enables risk differentiation between hospitals and specific professions.

## Methods

### Subjects and sampling

The Central Denmark Region is covering an area of 13,000 square kilometers. Seven hospitals are located in the region including Denmark’s largest hospital, Aarhus University Hospital, where more than half of all hospital staff in the region is employed. All healthcare workers and administrative personnel at the hospitals (including the pre-hospital services) and specialist practitioner clinics in the Central Denmark Region were invited by e-mail to be tested for antibodies against SARS-CoV-2. Blood-sampling was performed and organized by the Departments of Clinical Biochemistry at the hospitals. EDTA blood samples were collected from May 18 until June 19, 2020. The blood samples were transported to Aarhus University Hospital for centrifugation, and plasma was pipetted within 36 hours and stored at −30°C until analysis.

The SARS-CoV-2 prevalence in Denmark has been monitored nationally using reverse transcription polymerase chain reaction (RT-PCR) based viral RNA detection. The testing strategy has been adjusted several times since the outbreak (Figure 1, epidemic timeline). Healthcare workers have had easier access to testing than the general population, as they could be referred for testing by their employer. Until now (data from July 24) 1,057,333 individuals in Denmark have been tested, 13,392 are detected positive, and 612 individuals with coronavirus disease (COVID-19) have died (5).

**Figure 1.**
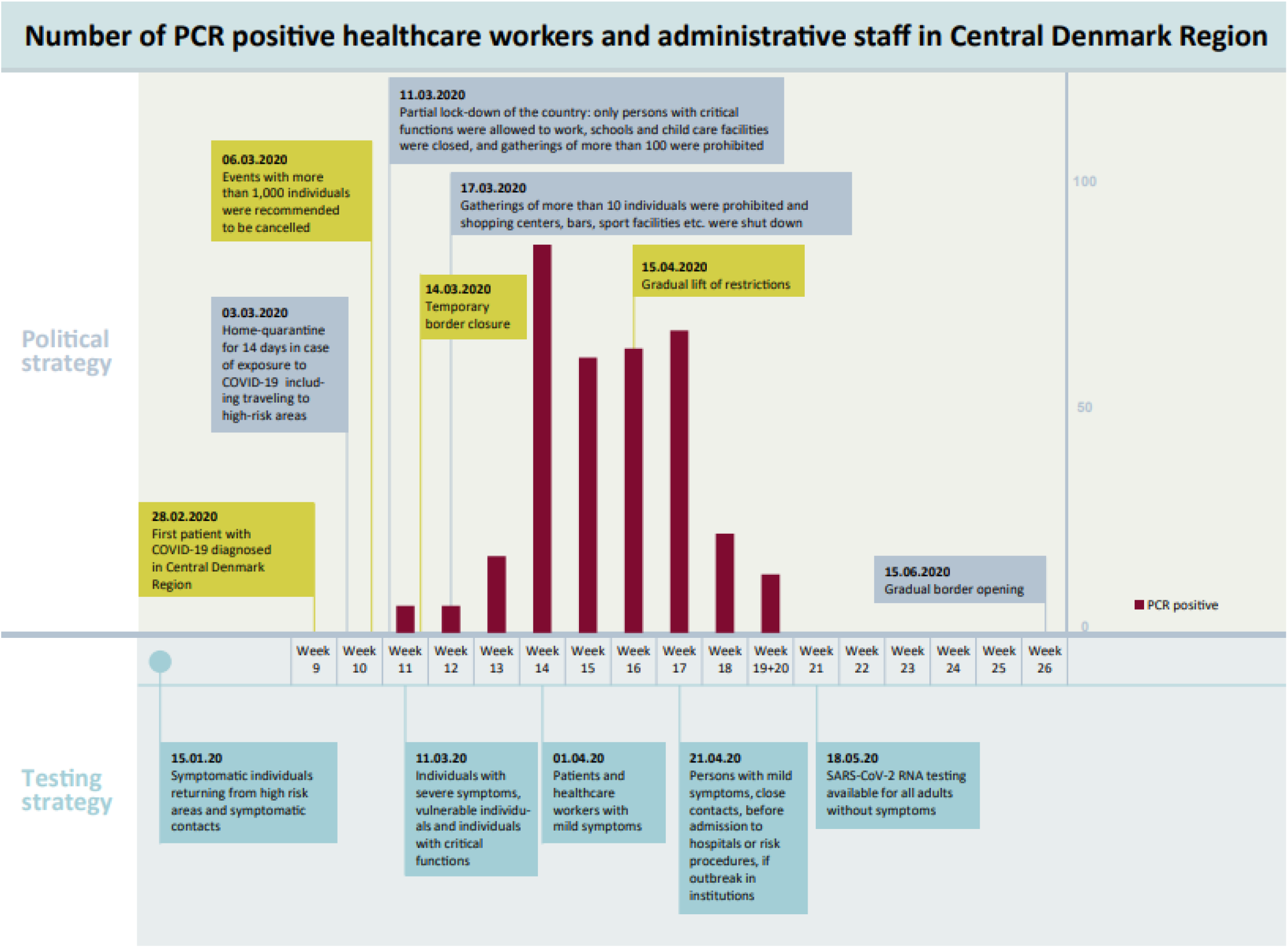
Epidemic timeline

### Serological testing

EDTA plasma was tested for antibodies to SARS-CoV-2 using a commercial SARS-CoV-2 total antibody enzyme-linked immunosorbent assay (ELISA, Wantai Biological Pharmacy Enterprise Co., Ltd., Beijing, China) according to the manufacturer’s instructions. The assay is based on a two-step incubation double-antigen sandwich principle that detects total antibodies in plasma binding SARS-CoV-2 spike protein receptor binding domain.

Results were based on a single test result. The sample absorbance (A) value was divided by a cutoff (CO) value for the ELISA plate based on an average absorbance value for 3 negative kit controls. A/CO values < 0.9 were considered negative, A/CO values = 0.9-1.1 were considered inconclusive, and A/CO values ≥ 1.1 were considered positive.

Performance characteristics of the Wantai SARS-CoV-2 total antibody ELISA has been determined in a Danish validation study (6). The assay had a sensitivity of 96.7% and a specificity of 99.5%. No cross-reactivity was observed.

Experienced staff at the Department of Clinical Immunology and the Department of Clinical Biochemistry, Aarhus University Hospital performed the tests. An internal PCR-confirmed SARS-CoV-2 positive control sample was tested at the beginning and in the end of each ELISA plate to ensure the accuracy of the analysis between departments, kits, and samples tested by one kit.

### PCR testing

In accordance with the national testing strategy, some of the healthcare- and administrative personnel participating in the study had previously been tested with RT-PCR technique in case of relevant COVID-19 symptoms or relevant risk of exposure. Due to changed national strategy for SARS-CoV-2 PCR testing during the pandemic, mainly persons who were severely ill or returned from a hotspot in Southern Europe or Asia were diagnosed using PCR prior to April 1, 2020. All PCR results were included, whether they had been referred for testing by their employer, general practitioner, or a hospital department.

PCR-analysis for SARS-CoV-2 RNA was performed in the Clinical Microbiology Department, either with Cobas® SARS-CoV-2 test on the Cobas® 6800 System with detection of the ORF-1a/b and E-gene or with in-house PCR analysis. For the in-house PCR-analysis, RNA was extracted from the samples and RT-PCR analysis was performed with the E gene assay from the Charité protocol (7) as recommended by WHO (8). Internal negative and positive controls were included in both the purification step and in the RT-PCR step.

### Risk groups and hotspots

Healthcare and administrative personnel demographic information, job title and workplace were obtained from the Central Denmark Region’s registration system of their employees. Healthcare workers at the hospitals were grouped according to their geographical location: 1) Herning and Holstebro Regional Hospitals serving the western part of the region, Regional Hospital West Jutland (RHWJ), 2) Viborg and Silkeborg Regional Hospitals serving the central part, Regional Hospital Central Jutland (RHCJ), and 3) Randers Regional Hospital, Horsens Regional Hospital, and Aarhus University Hospital servicing the eastern part (East). Psychiatry and social services, pharmacies, IT services, and administrative staff were grouped transregionally.

The seroprevalence among blood donors from the Central Denmark Region was also assessed. A total of 360 anonymized plasma samples from late June 2020 (180 from the western part and 180 from the eastern part of the region) were analyzed.

## Statistical analyses

Statistical analysis was performed in Stata/MP 16.1, RStudio 1.2, and R 3.6.0. Results were reported as percentages and percentage point (pp) differences with 95% confidence intervals (CIs). The Rogan Gladen estimator was used to estimate the true prevalence based on the estimates of the sensitivity and the specificity. To address both the population uncertainty and the uncertainties of the sensitivity and the specificity, percentile bootstrapping was used to make CIs, sampling the test results, sensitivity and specificity independently 10^8^ times each. For comparing two populations, the same methods were applied to obtain a set of 10^8^ samples of the estimated true prevalence for each population based on the same set of 10^8^ sensitivity and specificity estimates. The difference was assessed using a two-sided p-value, i.e. 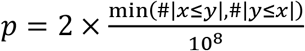. Predictors of risk were also analyzed by multivariable logistic regression analysis and presented as odds ratios (OR) with CIs.

## Ethics

This study is approved by the Danish Data Protection Agency (1-16-02-207-20) and by the Central Denmark Region. The regional scientific ethics committee of the Central Denmark Region concluded that this study did not require a scientific ethical approval (request no. 127 on ref. no. 110-72-1-20).

The SARS-CoV-2 antibody screening was performed at the request of the Danish Health Authority and the Danish Administrative Regions. Only consenting staff were tested and informed about their result.

## Results

A total of 25,950 healthcare workers and administrative personnel at hospitals, pre-hospital, and specialist practitioners in the Central Denmark Region were invited. Of these, 17,987 (69%) showed up for blood sampling, and 17,971 had samples available for SARS-CoV-2 antibody testing, see flowchart in Supplementary Figure 1.

The overall unadjusted seroprevalence was 3.7%, CI: 3.5%–4.0%. After adjusting for assay sensitivity and specificity including their CIs, the overall seroprevalence was 3.4%, CI: 2.5%–3.8%.

### Predictors of risk

There was no difference in seroprevalence according to sex. The youngest age group (younger than 30 years) had the highest seroprevalence, see Supplementary Table 1. The seroprevalence among hospital employees was higher in RHWJ than in RHCJ (11.9% vs 3.5%, difference = 8.5 pp, CI: 7.1–10.0) and East (11.9% vs 1.2%, difference = 10.7 pp, CI: 9.5–12.2), see Figure 2. Psychiatry and social services, pharmacies, and IT and administrative departments had a low adjusted seroprevalence (psychiatric departments: 1.0%, CI: 0.0%–1.8%; pharmacies, IT and administration departments: 1.7%, CI: 0.4%–3.0%).

**Figure 2.**
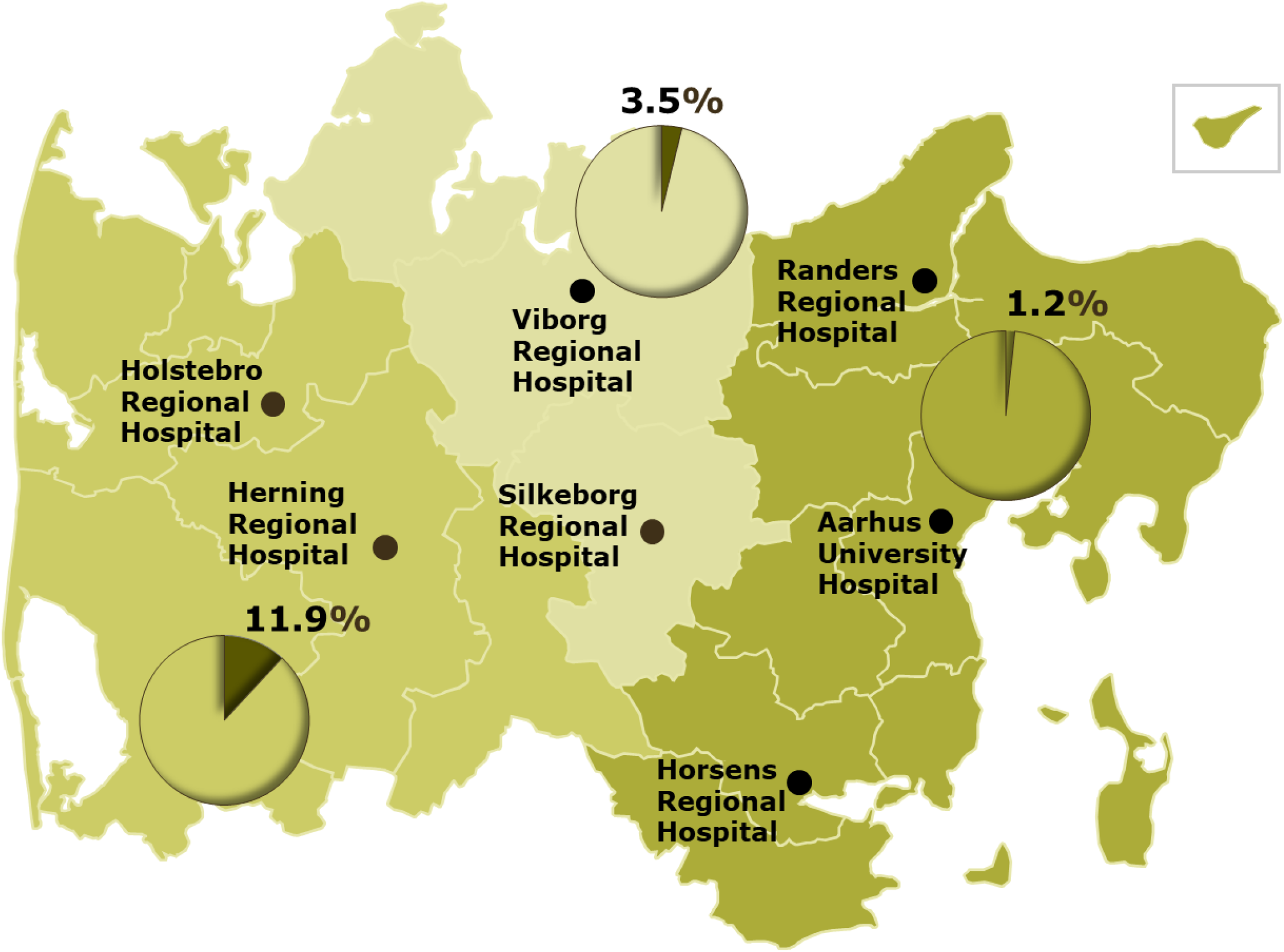
Distribution of adjusted seroprevalence according to geographical area in the Central Denmark Region. Herning and Holstebro Regional Hospitals are serving the western part; Viborg and Silkeborg Regional Hospitals are servicing the central part; Randers and Horsens Regional Hospitals, and Aarhus University Hospital are serving the eastern part.

The high seroprevalence in the RHWJ was analyzed separately. The emergency departments had the highest adjusted seroprevalence (29.7%) while departments with no or limited patient contact had the lowest seroprevalence (1.8%), see Table 2 for seroprevalence and pairwise comparisons of seroprevalence between groups. Risk of infection also depended on profession. In RHWJ, nursing staff (18.2%), medical doctors (12.8%), and biomedical laboratory scientists (12.9%) had higher seroprevalence compared to medical secretaries (2.5%), while no differences by profession were seen for RHCJ and East, see Table 3 for seroprevalence and Supplementary Table 2 for comparisons of seroprevalence according to profession.

**Table 1.** Age and sex stratified seroprevalence of anti-SARS-CoV-2.

| Age<br>years | Non-<br>reactive<br>n | Female |  |  | Non-<br>reactive<br>n | Male |  |  | Non-<br>reactive<br>n | Total |  |  |
| --- | --- | --- | --- | --- | --- | --- | --- | --- | --- | --- | --- | --- |
|  |  | n | Unadjusted % (CI) | Adjusted % (CI) |  | n | Unadjusted % (CI) | Adjusted % (CI) |  | n | Unadjusted % (CI) | Adjusted % (CI) |
| -29 | 1,669 | 97 | 5.49<br>(4.53-6.66) | 5.20<br>(3.89-6.46) | 247 | 19 | 7.14<br>(4.64-10.9) | 6.92<br>(4.09-10.8) | 1,916 | 116 | 5.71<br>(4.78-6.80) | 5.42<br>(4.09-6.59) |
| 30-39 | 3,189 | 115 | 3.48<br>(2.91-4.16) | 3.11<br>(2.00-3.87) | 605 | 29 | 4.57<br>(3.21-6.49) | 4.24<br>(2.52-6.21) | 3,794 | 144 | 3.66<br>(3.11-4.29) | 3.29<br>(2.20-4.02) |
| 40-49 | 3,971 | 159 | 3.85<br>(3.31-4.48) | 3.49<br>(2.40-4.22) | 589 | 16 | 2.64<br>(1.64-4.25) | 2.24<br>(0.81-3.88) | 4,560 | 175 | 3.70<br>(3.20-4.27) | 3.33<br>(2.26-4.01) |
| 50-59 | 3,980 | 143 | 3.47<br>(2.95-4.07) | 3.09<br>(2.02-3.79) | 525 | 18 | 3.31<br>(2.12-5.18) | 2.94<br>(1.35-4.84) | 4,505 | 161 | 3.45<br>(2.96-4.01) | 3.08<br>(2.01-3.74) |
| 60- | 2,041 | 60 | 2.86<br>(2.23-3.66) | 2.46<br>(1.31-3.32) | 464 | 12 | 2.52<br>(1.46-4.35) | 2.11<br>(0.64-3.97) | 2,505 | 72 | 2.79<br>(2.23-3.50) | 2.39<br>(1.28-3.18) |
| Total | 14,850 | 574 | 3.72<br>(3.43-4.03) | 3.36<br>(2.38-3.84) | 2,430 | 94 | 3.72<br>(3.05-4.54) | 3.36<br>(2.19-4.24) | 17,280 | 668 | 3.72<br>(3.45-4.01) | 3.36<br>(2.38-3.82) |

Inconclusive antibody results (n=23, 0.1%) were not included in this table.

**Table 2.** Distribution of seroprevalence according to affiliation in Regional Hospital West Jutland and pairwise comparisons of seroprevalence between groups (percentage point (pp) differences; significant differences after Bonferroni correction in bold).

|  | Emergency departments | Medical departments* | Surgical departments** | Department of Biochemistry | Service section*** | Anaesthesiology | Radiology and Nuclear Medicine | Departments with limited patient contact**** | Other***** | Total |
| --- | --- | --- | --- | --- | --- | --- | --- | --- | --- | --- |
| n | 165 | 829 | 556 | 150 | 286 | 300 | 153 | 362 | 125 | 2,926 |
| Adjusted % (CI) | 29.7<br>(23.1-37.6) | 15.0<br>(12.5-17.8) | 12.8<br>(10.0-16.0) | 12.0<br>(7.39-18.4) | 11.1<br>(7.68-15.5) | 9.89<br>(6.69-14.0) | 6.69<br>(3.54-12.4) | 1.79<br>(0.31-3.90) | 6.97<br>(3.31-13.1) | 12.0<br>(10.5-13.4) |
| Other | **22.8 (13.7-31.3)** | 8.07 (1.51-12.6) | 5.79 (-0.87-10.5) | 4.99 (-2.56-12.2) | 4.15 (-2.79-9.72) | 2.91 (-3.90-8.27) | -0.01 (-6.88-6.37) | -5.19 (-11.4;-1.07) | - |  |
| Departments with limited patient contact | **28.0 (21.0-35.9)** | **13.3 (10.1-16.3)** | **11.0 (7.60-14.4)** | **10.2 (5.20-16.7)** | **9.34 (5.40-13.8)** | 8.10 (4.38-12.3) | 5.18 (1.26-10.7) | - | - |  |
| Radiology and Nuclear Medicine | **22.8 (14.2-31.2)** | 8.08 (2.19-12.4) | 5.80 (-0.20-10.4) | 5.00 (-1.99-12.2) | 4.16 (-2.15-9.58) | 2.92 (-3.26-8.13) | - | - | - |  |
| Anaesthesiology | **19.9 (12.1-28.3)** | 5.15 (0.46-9.27) | 2.88 (-1.96-7.29) | 2.08 (-3.96-9.18) | 1.24 (-4.00-6.55) | - | - | - | - |  |
| Service section | **18.6 (10.7-27.1)** | 3.92 (-1.02-8.22) | 1.64 (-3.44-6.23) | 0.84 (-5.39-8.05) | - | - | - | - | - |  |
| Department of Biochemistry | **17.8 (8.55-26.8)** | 3.08 (-3.77-8.33) | 0.80 (-6.15-6.29) | - | - | - | - | - | - |  |
| Surgical departments | **17.0 (9.64-25.2)** | 2.28 (-1.67-6.08) | - | - | - | - | - | - | - |  |
| Medical departments | **14.7 (7.55-22.9)** | - | - | - | - | - | - | - | - |  |

\*Internal Medicine, Pediatrics, Oncology, and Neurology.
\*\*All surgical Departments, Obstetrics and Gynecology, Otorhinolaryngology, Head and Neck Surgery, and Ophthalmology.
\*\*\*Cleaning services, hospital porters, clothing and waste management, depot and archive, telephone switchboard, guidance for patients, relatives, and staff.
\*\*\*\*Occupational- and Social medicine, Physio- and occupational therapy, Administration, Department of Technical services, and Kitchen.
\*\*\*\*\*Participants without affiliation, e.g. students and participants on call, wage subsidy, and hourly waged.
Inconclusive results in the antibody test (n=4) were not included in this table.

The risk of infection was associated with workplace rather than place of living. The adjusted seroprevalence in participants working in RHWJ but living in the central or eastern part of the region was 10.7%, CI: 8.0%–13.7%), whereas the adjusted seroprevalence in participants living in the western part of the region, but working in East or RHCJ was 2.6% (CI: 0.7%–5.8%; difference: 8.1 pp, CI: 4.1–11.7).

To allow for multivariable analysis a logistic regression analysis was performed. In this analysis, however, we did not adjust for the assay sensitivity and specificity. The analysis confirmed that nursing staff, medical doctors, and biomedical laboratory scientists had a higher risk of testing positive than medical secretaries, who served as reference (OR = 7.3, CI: 3.5–14.9; OR = 4.0, CI: 1.8–8.9; and OR = 5.0, CI: 2.1–11.6, respectively) while adjusting for age group and sex. The analysis also showed that individuals younger than 30 had a higher risk of testing positive for SARS-CoV-2 antibodies than the other age groups combined (OR = 1.9, CI: 1.4–2.6). The analysis showed no effect of sex.

### Seroprevalence among blood donors

The adjusted seroprevalence among 360 blood donors was low in both the western part (1.2%, CI: 0.0%–4.4%) and the eastern part of the region (0.6%, CI: 0.0%–3.5%; difference = 0.6 pp, CI: −2.4–3.6).

### Association between SARS-CoV-2 RNA and total antibodies

During February 28 to June 23, 2020, 4,803 (26.7%) of the participants in the seroprevalence initiative (n = 17,971) had been tested for SARS-CoV-2 RNA in an oro- or nasopharyngeal swab or tracheal aspirate by RT-PCR, see Table 4. A total of 341 (7.1%) tested positive for SARS-CoV-2 RNA, and among these, 334 (98.0%) were subsequently seropositive. Among the total 668 seropositive participants, 433 (64.8%) had been tested for viral RNA and 50.0% had a positive result.

**Table 3.** Distribution of seroprevalence according to profession in hospital employees in the Central Denmark Region (n=15,261).

|  |  | Central Denmark Region |  |  |  |  |
| --- | --- | --- | --- | --- | --- | --- |
|  |  | Nursing staff * | Medical doctors | Biomedical<br>Laboratory<br>Scientists | Medical<br>secretaries | Other |
| RHWJ | Adjusted % (CI) | 18.2 (15.9-20.7) | 12.8 (9.67-16.6) | 12.9 (8.21-19.4) | 2.52 (0.74-5.31) | 5.77 (4.03–7.62) |
| RHCJ | Adjusted % (CI) | 5.22 (3.76-6.60) | 2.99 (1.21-5.47) | 2.01 (0.21-5.71) | 1.57 (0.10-3.89) | 1.62 (0.37–2.91) |
| East | Adjusted % (CI) | 1.24 (0.22-1.78) | 1.89 (0.70-2.91) | 1.43 (0.18-2.75) | 1.09 (0.00-2.26) | 0.80 (0.00–1.45) |
| Total (n) |  | 7,011 | 2,066 | 1,017 | 1,351 | 3,796 |

\*Nurses, social- and healthcare assistants, and radiographers.
RHWJ: Regional Hospital West Jutland
RHCJ: Regional Hospital Central Jutland
East: Randers and Horsens Regional Hospital, and Aarhus University Hospital serving the eastern part
Inconclusive results in the antibody test (n=20) were not included in this table.

**Table 4.** Association between SARS-CoV-2 RNA by RT-PCR and total antibodies.

|  | SARS-CoV-2 total antibodies |  |  |  |  |
| --- | --- | --- | --- | --- | --- |
| SARS-CoV-2<br>PCR | Negative |  | Positive |  |  |
|  | n | % | n | % |  |
| Negative | 4,358 | 97.8 | 99 | 2.2 | 4,457 |
| Positive | 6 | 1.8 | 334 | 98.2 | 340 |
|  | 4,364 |  | 433 |  | 4,797* |

\*Inconclusive results in the antibody test (n=6) were not included in this table.

Only 23.1% of the staff tested for SARS-CoV-2 RNA were employed in RHWJ, but they accounted for 66.6% of the SARS-CoV-2 RNA positive participants. Among 351 seropositive participants employed at RHWJ, 270 were at some point additionally tested for viral RNA and 224 tested positive for SARS-CoV-2 RNA (63.8% of seropositives).

## Discussion

The adjusted seroprevalence of antibodies to SARS-CoV-2 in healthcare workers and administrative personnel at hospitals, pre-hospital services, and specialist practitioners in the Central Denmark Region was 3.4%. There were, however, sizable differences in seroprevalence between hospitals ranging from less than 2% in East to almost 12% in the RHWJ, even though the distance between the hospitals situated furthest apart is only 120 km. In RHWJ, the risk was highest in the emergency departments and higher in departments and professions with frequent patient contact, while no such pattern was seen in the central or eastern parts.

Among the seropositive staff, 65% had previously been tested for SARS-CoV-2 RNA, and 50% of the seropositives had already been confirmed SARS-CoV-2 RNA positive. This percentage was particularly high in the RHWJ where 64% of all seropositives had a prior positive test for SARS-CoV-2 RNA. This indicates that personnel suspected of COVID-19, to a high degree, are being referred for PCR testing. We would expect to have found a higher percentage of concomitant seropositive and PCR positive staff if the early test strategies (prior to April 1, 2020) had allowed for PCR testing of asymptomatic or mildly symptomatic personnel. The combination of serological and molecular findings also allowed us to verify the sensitivity of the serological assay used: 98% of employees previously tested positive for viral RNA had a positive test for SARS-CoV-2 antibodies.

Large differences in seroprevalence in healthcare workers have been reported (1-43%) (1, 2, 9) similar to the large differences in severity of the COVID-19 epidemic between countries (10), however comparison to the background population level has not been reported.

In Denmark, the seroprevalence in blood donors differs between areas with low prevalence in the Central Denmark Region (11). However, similar to findings from Italy large geographical differences within the Central Denmark Region have been shown in this study but within an even smaller geographical area (1, 4). While there is evidence of a high number of infected individuals during March and April among staff working at RHWJ and a higher incidence of infection in patients from the hospital’s service area (10), this does not translate into a high seroprevalence in the background population of the area: First, the seroprevalence in blood donors from the area tested during April was 1% (11). Second, the seroprevalence in 180 anonymized blood donations given in June in the RHWJ service area was 1.2%. Third, the seroprevalence was low in departments and professions with no patient contact. Fourth, we found that staff living but not working in the western part of the region had a low seroprevalence while staff working but not living in the western part of the region had a high prevalence of antibodies suggesting in-hospital transmission.

A recently published study among health-care workers in the Capital Region of Denmark reported a similar seroprevalence of 4.0% (12). However, we found much greater differences between hospitals and departments. Thus, we report a seroprevalence of 29.7% among staff in the emergency departments in RHWJ whereas a seroprevalence of 7.2% was reported from dedicated COVID-19 wards in Copenhagen despite a high level of population transmission in the Capital Region of Denmark (10).

Several reasons for the discrepancy may exist. We used an ELISA with a high sensitivity of 96.7%, which was run in two centralized laboratories by experience staff. In the Capital Region, point-of-care tests with a lower sensitivity (87.2%) were distributed to the departments and results were read and reported by the individual. Whereas we detected antibodies in 98% of individuals previously tested positive for viral RNA this was only the case for 64.2% in the study from the Capital Region.

Possible explanations for transmission of SARS-CoV-2 to the staff in RHWJ are higher levels of population transmission in the service area of RHWJ (cumulative incidence of 210 pr. 100.000 inhabitants vs. 85 pr. 100.000 in the eastern part) (10). Moreover, older hospital buildings with less space, less single-bed rooms, and less optimal facilities for isolation of patients with infectious diseases may have added to the risk. Whether inadequate access to proper personal protective equipment (PPE) and/or insufficient training may also play a role cannot be answered by this study. Another possible explanation for a higher risk of in-hospital transmission could be that the COVID-19 patients in the western part of the region were older (data not shown). Therefore, they may have required more help from the healthcare workers and consequently more frequent and closer contact. Furthermore, the older age of the patients may have obscured the clinical symptoms leading to less testing for SARS-CoV-2 in this age group. This in contrast to e.g. skiing vacation with known risk of exposure, which was common among younger individuals infected early in the epidemic in Denmark (13). In the eastern part of the region, contact tracing and testing was performed more aggressively at Aarhus University Hospital early in the epidemic, which could have reduced the burden of disease.

Studies have shown that risk factors for COVID-19 among healthcare workers include working at a clinical department, working in a high risk versus general department, suboptimal hand hygiene before or after patient contact, longer work hours, improper PPE use, working as a medical doctor, contact with COVID-19 patients, contact with super-spreader patients, and staff of younger age developing more severe disease maybe as a sign of more intense exposure (4, 14–17). In line with this, we found that younger staff were more likely to be seropositive. Frequent shifts and closer contact to newly admitted and yet undiagnosed patients among young employees and staff working at the emergency department may lead to higher risk of exposure. The high risk of infection among biomedical laboratory scientists reflects that in Denmark this group of staff has frequent patient contact since they are responsible for drawing blood.

Half of the staff with a positive serological test had been tested positive by PCR prior to the antibody testing. Since 30-40% of COVID-19 patients may be asymptomatic (18) this indicates that a thorough and targeted testing activity has been performed. However, it also raises the case whether healthcare workers should be screened for SARS-CoV-2 on a regular basis since transmission can occur even in the absence of symptoms (19–21).

### Strengths and limitations

This is one of the largest studies assessing the prevalence of SARS-CoV-2 antibodies among healthcare workers to date, and the study was performed with an assay with a proven sensitivity of 97%. The participation rate was 69%. Yet, health care workers not able to work e.g. due to sickness leave, maternity leave, etc. were also invited to participate; however, not expected to be tested. Participation may have depended on exposure or suspicion of infection (Supplementary Figure 1). On the other hand, healthcare workers that had already been diagnosed with COVID-19 may have been less likely to participate since they were expecting to test positive. We may therefore, either over- or underestimate the true prevalence. Information about original job title and workplace was retrieved from the employer’s registration system. However, due to the epidemic and subsequent closing or partly closing of some departments, some employees were transferred to departments treating COVID-19 patients. Since information about use of PPE or specific tasks were not available, more detailed information about risk factors could not be assessed.

This study was done after the epidemic had slowed down in Denmark in a time period with few new infections. The median time from symptom onset to detection of total SARS-CoV-2 antibodies is 11 days (22) meaning that most infected staff would already have seroconverted when this survey was done. The dynamics of antibody concentrations and whether they wane over time is still unknown, but from our own experience with a small group (n = 12) of convalescent plasma donors, serological test results and virus neutralizing antibody titers remain unchanged during three months of follow-up. Among PCR-confirmed COVID-19 cases, 98% were antibody-positive individuals, which is consistent with the results of the validation of the assay. Even though the specificity of the Wantai assay was acceptable at 99.5%, the low seroprevalence in most hospitals implies a low positive predictive value of the test. Since we do not have a Gold Standard to confirm positive or negative results, we estimated the confidence interval with a method that took both the sample variation and the uncertainty in the sensitivity and specificity into account adjusting for the test performance. Finally, cross-reactivity with e.g. other coronaviruses could result in false-positive antibody test results.

Taking these limitations into account, this study should raise awareness of means to avoid in-hospital transmission by improving institutional infection control measures including training on infection control procedures and ensuring compliance with PPE use (23) as well as considering testing of both symptomatic and asymptomatic healthcare workers for SARS-CoV-2 on a regular basis (21, 24).

## Conclusion

In conclusion, we found large intraregional differences in the prevalence of SARS-CoV-2 antibodies among staff working in the healthcare sector within a small geographical area of Denmark. The seroprevalence in the western part of the region was significantly higher among healthcare workers with patient contact than among the background population suggesting in-hospital transmission. Half of all seropositive staff had already been tested positive by PCR prior to this survey indicating a targeted testing strategy but also highlighting a need for PCR test screening in healthcare workers.

## Data Availability

Data available on request.

## Author contributions

SJ, SM, TG, KN, LØ, MKT, HJM and CE planned the study. SJ, SM, AMA, JKB, JDR, TG and CE analyzed and interpreted the data. SJ, SM, CE and KAK drafted the manuscript. SM, CE, TG, MKT and HJM were responsible for the laboratory analyses. All authors were involved in critically revising the manuscript and approved the final version.

## Acknowledgements

We thank all the involved staff from the Department of Clinical Immunology, Department of Clinical Biochemistry, Department of Clinical Microbiology at Aarhus University Hospital, as well as staff from the Department of Clinical Biochemistry at Regional Hospital West Jutland, Regional Hospital Central Jutland, Horsens, Randers Hospital, and Grenå Sundhedshus for their exceptional work in coordinating the study and technical assistance.

## Supplementary

**Supplementary Figure 1.**
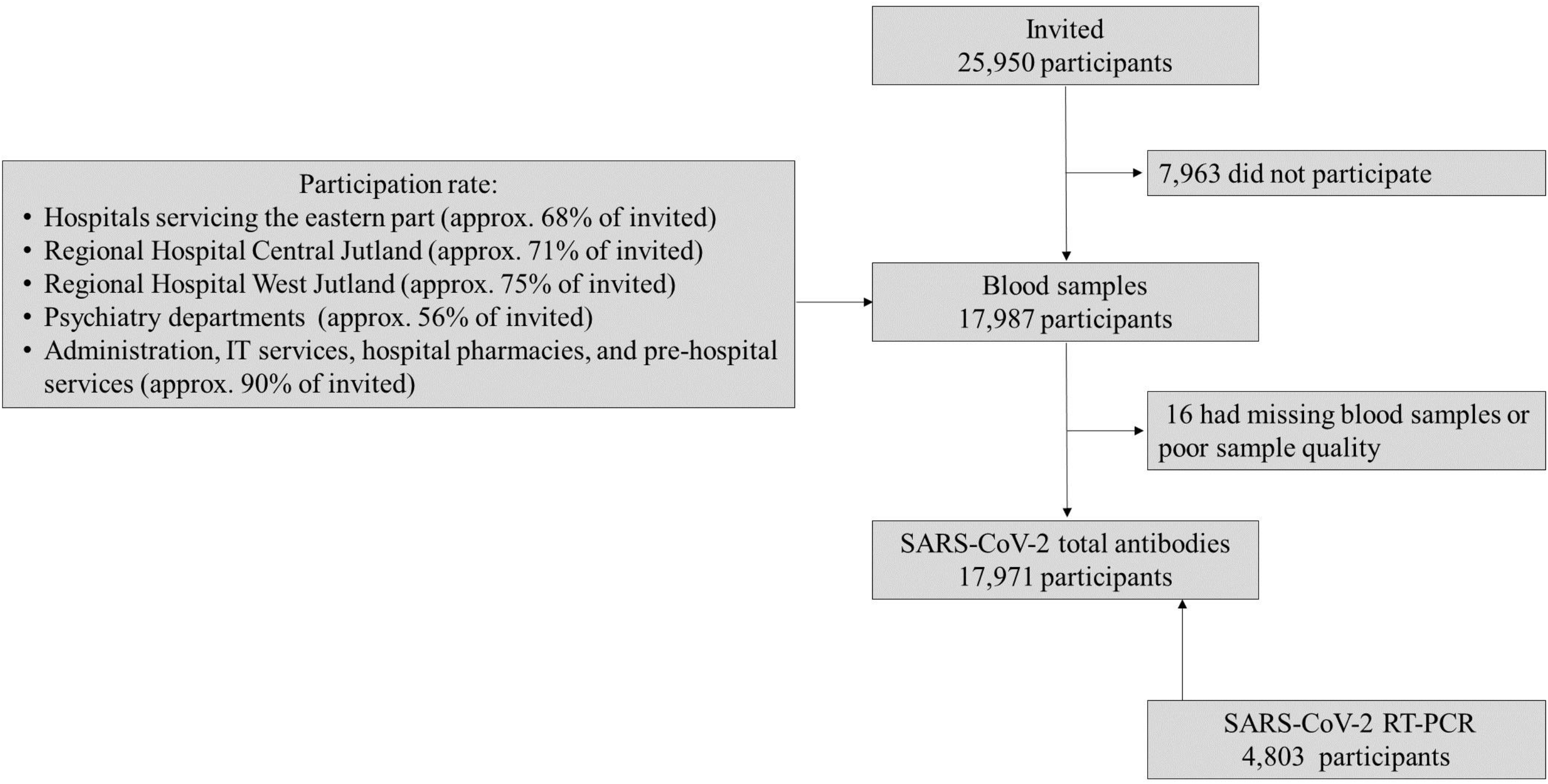
Flowchart

**Supplementary Table 1.**
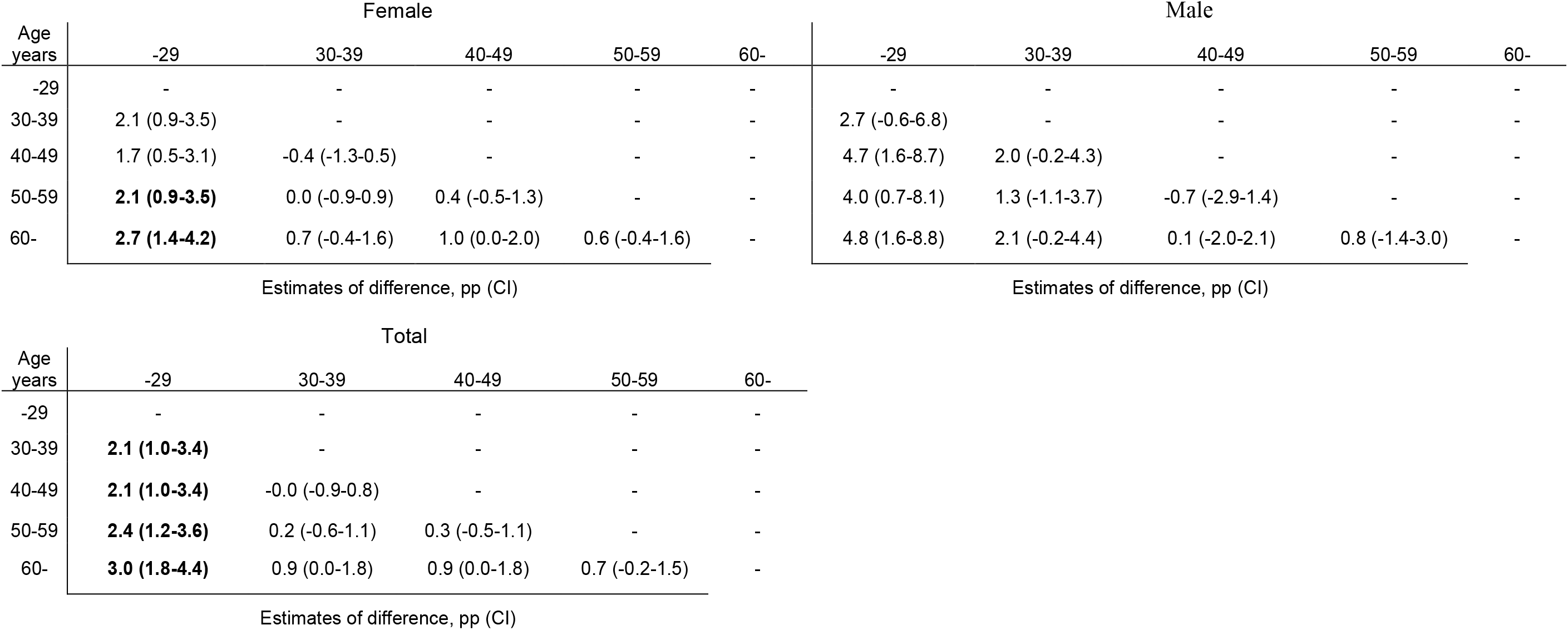
Pairwise comparisons of seroprevalence between age strata. Percentage point (pp) differences were calculated by subtracting the seroprevalence in the older age category from the seroprevalence in the younger; significant differences after Bonferroni correction in bold.

| Age<br>years | Female |  |  |  |  | Male |  |  |  |  |
| --- | --- | --- | --- | --- | --- | --- | --- | --- | --- | --- |
|  | -29 | 30-39 | 40-49 | 50-59 | 60- | -29 | 30-39 | 40-49 | 50-59 | 60- |
| -29 | - | - | - | - | - | - | - | - | - | - |
| 30-39 | 2.1 (0.9-3.5) | - | - | - | - | 2.7 (-0.6-6.8) | - | - | - | - |
| 40-49 | 1.7 (0.5-3.1) | -0.4 (-1.3-0.5) | - | - | - | 4.7 (1.6-8.7) | 2.0 (-0.2-4.3) | - | - | - |
| 50-59 | **2.1 (0.9-3.5)** | 0.0 (-0.9-0.9) | 0.4 (-0.5-1.3) | - | - | 4.0 (0.7-8.1) | 1.3 (-1.1-3.7) | -0.7 (-2.9-1.4) | - | - |
| 60- | **2.7 (1.4-4.2)** | 0.7 (-0.4-1.6) | 1.0 (0.0-2.0) | 0.6 (-0.4-1.6) | - | 4.8 (1.6-8.8) | 2.1 (-0.2-4.4) | 0.1 (-2.0-2.1) | 0.8 (-1.4-3.0) | - |
| Estimates of difference, pp (CI) |  |  |  |  | Estimates of difference, pp (CI) |  |  |  |  |  |
| Age<br>years | Total |  |  |  |  |  |  |  |  |  |
|  | -29 | 30-39 | 40-49 | 50-59 | 60- |  |  |  |  |  |
| -29 | - | - | - | - | - |  |  |  |  |  |
| 30-39 | **2.1 (1.0-3.4)** | - | - | - | - |  |  |  |  |  |
| 40-49 | **2.1 (1.0-3.4)** | -0.0 (-0.9-0.8) | - | - | - |  |  |  |  |  |
| 50-59 | **2.4 (1.2-3.6)** | 0.2 (-0.6-1.1) | 0.3 (-0.5-1.1) | - | - |  |  |  |  |  |
| 60- | **3.0 (1.8-4.4)** | 0.9 (0.0-1.8) | 0.9 (0.0-1.8) | 0.7 (-0.2-1.5) | - |  |  |  |  |  |
| Estimates of difference, pp (CI) |  |  |  |  |  |  |  |  |  |  |

**Supplementary Table 2.**
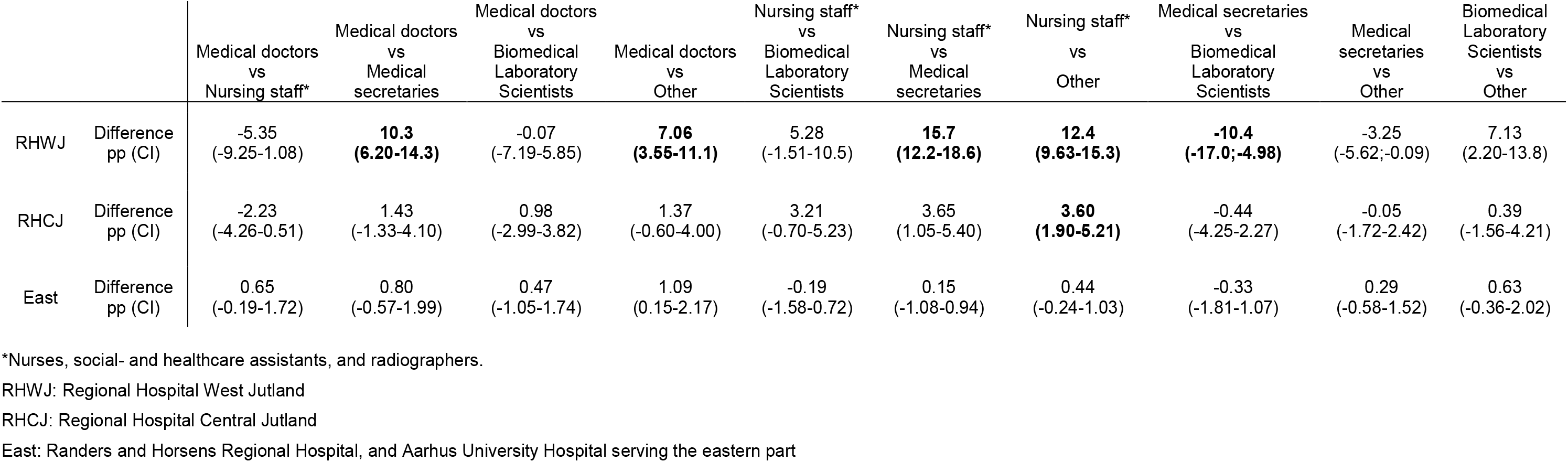
Differences in seroprevalence between professions in the Central Denmark Region (percentage point (pp) differences; significant differences after Bonferroni correction in bold).

|  |  | Medical doctors<br>vs<br>Nursing staff* | Medical doctors<br>vs<br>Medical<br>secretaries | Medical doctors<br>vs<br>Biomedical<br>Laboratory<br>Scientists | Medical doctors<br>vs<br>Other | Nursing staff*<br>vs<br>Biomedical<br>Laboratory<br>Scientists | Nursing staff*<br>vs<br>Medical<br>secretaries | Nursing staff*<br>vs<br>Other | Medical secretaries<br>vs<br>Biomedical<br>Laboratory<br>Scientists | Medical<br>secretaries<br>vs<br>Other | Biomedical<br>Laboratory<br>Scientists<br>vs<br>Other |
| --- | --- | --- | --- | --- | --- | --- | --- | --- | --- | --- | --- |
| RHWJ | Difference<br>pp (CI) | -5.35<br>(-9.25-1.08) | **10.3**<br>**(6.20-14.3)** | -0.07<br>(-7.19-5.85) | **7.06**<br>**(3.55-11.1)** | 5.28<br>(-1.51-10.5) | **15.7**<br>**(12.2-18.6)** | **12.4**<br>**(9.63-15.3)** | **-10.4**<br>**(-17.0;-4.98)** | -3.25<br>(-5.62;-0.09) | 7.13<br>(2.20-13.8) |
| RHCJ | Difference<br>pp (CI) | -2.23<br>(-4.26-0.51) | 1.43<br>(-1.33-4.10) | 0.98<br>(-2.99-3.82) | 1.37<br>(-0.60-4.00) | 3.21<br>(-0.70-5.23) | 3.65<br>(1.05-5.40) | **3.60**<br>**(1.90-5.21)** | -0.44<br>(-4.25-2.27) | -0.05<br>(-1.72-2.42) | 0.39<br>(-1.56-4.21) |
| East | Difference<br>pp (CI) | 0.65<br>(-0.19-1.72) | 0.80<br>(-0.57-1.99) | 0.47<br>(-1.05-1.74) | 1.09<br>(0.15-2.17) | -0.19<br>(-1.58-0.72) | 0.15<br>(-1.08-0.94) | 0.44<br>(-0.24-1.03) | -0.33<br>(-1.81-1.07) | 0.29<br>(-0.58-1.52) | 0.63<br>(-0.36-2.02) |

\*Nurses, social- and healthcare assistants, and radiographers.
RHWJ: Regional Hospital West Jutland
RHCJ: Regional Hospital Central Jutland
East: Randers and Horsens Regional Hospital, and Aarhus University Hospital serving the eastern part

